# Economic burden of smoking attributed illnesses in Pakistan

**DOI:** 10.1101/2020.06.15.20131425

**Authors:** Muhammad Arif Nadeem Saqib, Ashar Malik, Ibrar Rafique, Faiz Ahmed Raza, Obaidullah, Shaimuna Fareeha Sajjad, Sumera Abid, Rukhsana Majid, Tahira Kamal, Ziauddin Islam

**Author notes:** Corresponding author: Dr. Muhammad Arif Nadeem Saqib, Senior Research Officer, Pakistan Health Research Council, Sector G-5/2, Constitution Avenue, Sharah-e-Jamhuriat, Islamabad, Email;, 051-9207368.

## Abstract

**Background:** The estimates of economic burden due to smoking attributed illnesses provide an opportunity to assess its overall impact on the economy and generate evidence for public health policy interventions for tobacco control. In this study, we estimated out of pocket expenditures on tobacco attributed illnesses and smoking attributable burden in Pakistan.

**Methods:** We used a prevalence-based disease-specific cost approach by including three major tobacco attributed illnesses i.e. lung cancer, chronic obstructive pulmonary disease, and cardiovascular diseases. Our analysis included out of pocket healthcare expenditures including direct and indirect costs which were estimated by interviewing the patients of selected illnesses. The smoking-attributable expenditure was calculated by the WHO tool kit.

**Results:** In 2018, the economic burden attributed to smoking related illnesses was Rs 192 billion (USD 1.3 billion). Smoking-attributable expenditure on cardiovascular disease was Rs 123 billion (USD 0.9 billion) which was 69% of the total economic cost of tobacco attributed illnesses in Pakistan. The economic cost in males was nearly three times higher than females.

**Conclusions:** Our study showed a significant economic burden due to tobacco attributed illnesses in Pakistan which can be prevented by implementing tobacco control policies effectively.

## MAIN TEXT

### Introduction

Tobacco use is an important behavioral problem and has harmful health consequences like chronic obstructive pulmonary disease (COPD), cardiovascular diseases (CVDs) and lung cancers ^1^. According to World Health Organization (WHO), tobacco use is responsible for the death of one in ten adults’ worldwide (about 5 million deaths each year) and millions develop tobacco attributed illnesses resulting in chronic disability ^2^. These tobacco attributed illnesses have a huge economic impact on the individuals, families and societies. Studies to assess the economic burden of tobacco usage have been carried out in high income countries like the United Kingdom, Canada, Australia as well as in developing countries like China, Vietnam, India and Bangladesh ^3-7^. The estimates of the costs attributed to tobacco use and its associated illnesses are valuable to policy-makers, particularly in planning health service provision and other items of public expenditure.

Tobacco consumption is common in Pakistan where 19.1% of adults used tobacco products including 12.4% smoker and 7.7% smokeless tobacco users ^8^. Similarly, 10.4% of the youth used tobacco ^9^. Though sufficient data is available about the prevalence of tobacco in the country, little is known about the impact of tobacco consumption on an individual’s health, its economic burden on the patients and the health care system. We planned this study to estimate the out of pocket expenditure incurred by the patients on treatment of tobacco attributed illnesses and to determine the economic burden of smoking attributed illnesses in Pakistan.

## Methods

We used a prevalence-based attributable-risk approach which measures the value of resources used (direct costs) or lost (indirect costs) from tobacco attributed illnesses and deaths in a given period, regardless of the time of tobacco use and the onset of the disease. This approach helps to estimate the overall economic burden due to tobacco use on society. The direct cost includes consultation fees, cost of medicine, hospitalization, and transportation to health centers while and indirect cost consists of the number of workdays missed by the patient and attendants (income loss) and premature deaths. Tobacco attributed illnesses included in this study were COPD, CVDs and CA lung.

Primary data was collected during the calendar year 2017-2018 (June 2017 to October 2018) for estimation of out of pocket expenditure on tobacco attributed illnesses. We selected 14 hospitals (06 public and 08 private) based on their level of health care services (tertiary care) and the geographical location (located at provincial and federal capitals namely Peshawar, Lahore, Quetta, Karachi and Islamabad) to represent the entire country. A structured questionnaire with closed ended questions was administered to the patients/their attendants who had been seeking care at the sampled hospitals. Data pertained to the basic demographics of the patient and their attendants, disease profile, type and episodes of care availed. This followed on the socioeconomic status of the patient/families. Data on health seeking included questions on expenditures relating to consultation, medicines, laboratory and radiological examinations, overnight stay, food and travel of the patient and attendant.

A total of 1549 patients were enrolled including 40.3% COPD, 30.9% CVDs and 19.6 % CA lungs while the rest were other illnesses (oral cancer etc). Those who had reported to currently smoking were 50.7% including 36.7% male current smokers. The data of persons aged 35 years and older were analyzed keeping in view high relative risk of developing COPD, CVD and CA Lungs in this age group. The final sample for analysis of those aged 35 years and above who were smoking at the time and had reported any of the three selected illnesses was 44% of the total sample (n=688). The detailed characteristics of the participants are given in table 1.

**Table 1:** Demographic characteristic of the study participants

| **Description** | **All (n=1549)** | **Aged ≥35 and current smokers (n=688)** |
| --- | --- | --- |
| **Mean Age** | 56.93 (±15.29) | 60.3 (±12.6) |
| **Gender** |  |  |
| Male | 1168 (75.4%) | 528 (76.7%) |
| Female | 381 (24.6%) | 160 (23.3%) |
| **Education** |  |  |
| Illiterate | 820 (52.9%) | 408 (59.3%) |
| Primary or less | 200 (12.9%) | 82 (12.0%) |
| Secondary or Less | 378 (24.5%) | 161 (23.3%) |
| High School and Above | 151 (9.8%) | 37 (5.4%) |
| **Tobacco attributed diseases** |  |  |
| CVDs | 624 (40.3%) | 282 (41.0%) |
| COPD | 607 (39.2%) | 262 (38.1%) |
| CA Lungs | 304 (19.6%) | 144 (20.9%) |

CA Lungs: Lung cancer, COPD: Chronic obstructive pulmonary disease, CVDs: Coronary vascular diseases

Costs were stratified by gender, type of disease and type of costs i.e. direct and indirect cost. The direct cost includes the healthcare i.e. cost of the consultation, hospital stay, diagnosis (radiological and laboratory), surgical or other procedures, medicines and equipment while other costs were the cost of food and diet, travel of the patient and caregiver for seeking care. The indirect cost includes the productively losses of patients and attendants including time spent on visiting the hospital and stay in the hospital while seeking care. All costs were estimated using the median and inter-quartile range.

The literature search was carried out on Google scholar and Pubmed in the months of June-July 2019 using the search terms, “Prevalence”, “Incidence”, Lung cancer, Chronic Pulmonary Obstructive Disease, Cardiovascular disease, Stroke, “Ischemic heart attack”, “Myocardial infarction”, “Pakistan”. Later on, the search strategy included “South Asia”, “India”, “Bangladesh”, “Sri Lanka” and “Afghanistan”. The literature search was restricted to articles published after 2000 in English language only. The literature search was further extended to expert in the respective clinical expertise such as cardiologist, cardiothoracic surgeons, pulmonologist, oncologists and epidemiologist to know about the published reports and literature on prevalence COPD, CVDs and CA Lungs in Pakistan or neighboring/ regional countries.

Based on the literature search, the CA lungs estimates were calculated using overall Cancer Incidence in Pakistan (111.8 per 100,000 people) and the proportion of lung cancers (4.2% of all cases) ^10^. The prevalence of COPD and CVDs were taken as 2.1 % and 17.5% respectively^11 12^. Tobacco attributed mortality data were obtained from the World Health Organization estimates on death rates and their proportion attributable to tobacco for COPD, CVDs and CA lungs ^13^. The overall prevalence and relative risk of mortality due to COPD, CVDs and CA lungs used for calculation are given in table 2.

**Table 2:** Overall prevalence and relative risk of mortality due to CA lung, COPD and CVDs

| **Description** | **Male** | **Female** | **Total** |
| --- | --- | --- | --- |
| **Prevalence to tobacco attributed diseases** |  |  |  |
| CA Lungs | 0.007 | 0.003 | 0.005 |
| COPD | 0.021 | 0.021 | 0.021 |
| CVDs | 0.166 | 0.183 | 0.175 |
| **Death Rate** |  |  |  |
| CA Lungs | 364 | 6 | 22 |
| COPD | 1153 | 95 | 103 |
| CVDs | 8653 | 560 | 585 |
| **Death rate attributable to tobacco (per 100000)** |  |  |  |
| CA Lungs | 322 | 0 | 15 |
| COPD | 847 | 4 | 38 |
| CVDs | 133 | 1 | 35 |
| **Proportion of deaths attributable to tobacco (%)** |  |  |  |
| CA Lungs | 88 | 3 | 17 |
| COPD | 73 | 5 | 37 |
| CVDs | 2 | 0 | 6 |

Source WHO2012: CA Lungs: Lung cancer, COPD: Chronic obstructive pulmonary disease, CVDs: Coronary vascular diseases

Calculation of smoking impact ratio (SIR) and smoking attributed fraction (SAF) was based on these estimates using estimation methods suggested by WHO ^14^. SIR was estimated using gender specific mortality attributable to specific COPD, CVDs and CA lungs. SAF was estimated using the gender specific and illnesses specific SIR and the number of 35+ population affected by the respective illness. The tobacco attributed expenditure (TAE) was estimated by multiplying SAF for each selected disease with the yearly cost for treatment of the respective disease.

Data were entered, cleaned and coded into MS Excel. All analysis was carried out using STATA version 15.1. The costs and economic burden were estimated in Pakistan rupees (Rs) and converted to US$ exchange rate with Rs in 2018 i.e. 1 USD equal to 139.80 Rs.

The study was conducted as per Helsinki declaration. The Ethical Clearance was obtained from Institutional Bioethics Committee of Pakistan Health Research Council, Islamabad. Written informed consent was taken from participants prior to enrolment in the study.

## Results

Overall the annual out of pocket expenditures of tobacco attributed illnesses was Rs. 42566 (USD 304.47). Healthcare cost constituted the largest share (>60%) in the total cost. The expenditure on the management of CA lungs was high i.e. Rs 128425 (USD 918.63) followed by COPD and CVDs i.e. Rs. 39038 (USD 279.24) and Rs 30640 (USD 219.17) respectively Cost estimates showed high variability in the case of CVDs where the estimates of IQR were higher than the median costs (Table 3). It was observed that the out of pocket expenditures for the management of male respondents was higher than female.

**Table 3:** Average direct, indirect and total costs (median) on the management of CA lungs, COPD and CVDs for the year 2018

| **Illness** | **Currency** | **Health Care Cost** | **Other costs** | **Productivity losses** | **Total costs** |
| --- | --- | --- | --- | --- | --- |
| CA Lungs | Rs. | 70125(79200) | 40400 (76540) | 10500 (15850) | 128425 (173300) |
|  | USD | 501.6 (566.52) | 288.98 (547.50) | 75.10 (113.38) | 918.63 (1239.63) |
| COPD | Rs. | 21000(39099) | 4420 (7800) | 11600 (12000) | 39037.5 (55925) |
|  | USD | 150.21(279.68) | 31.61 (55.79) | 82.97 (85.84) | 279.23 (400.04) |
| CVDs | Rs. | 17804(45000) | 4000 (6100) | 7500 (19420) | 30640 (68046) |
|  | USD | 127.35(321.89) | 28.61(43.63) | 53.64 (138.91) | 219.17 (486.74) |
| Total | Rs. | 27000(58560) | 5000 (12100) | 10000 (15200) | 42566 (93074) |
|  | USD | 193.13(418.88) | 35.76 (86.55) | 71.53 (108.73) | 304.47 (665.77) |

Inter quartile range in parenthesis, CA Lungs: Lung cancer, COPD: Chronic obstructive pulmonary disease, CVDs: Cardiovascular diseases

Table 4 shows the SIR and SAF of tobacco use by gender and illnesses as well as the tobacco attributed economic burden. Overall there were minimal gender related differences in SIR or SAF except for COPD. The COPDs and CVDs had much higher disease specific SAF i.e. 36.66% and 30.40% respectively in male as compared to the CA lung. The total tobacco attributed the economic burden of the selected three illnesses was Rs 192 billion (USD 1.37 billion) which was significantly higher for male. Similarly, within the illnesses, the burden attributed to CVDs constituted the largest share i.e. 64% (Table 4).

**Table 4:** Smoking impact ratio, smoking attributable fraction and tobacco attributed economic burden (aged >35 years) in 2018

| Description | Male |  |  | Female |  |  | Total (Rs) |
| --- | --- | --- | --- | --- | --- | --- | --- |
|  | SIR | SAF | Cost (Rs) | SIR | SAF | Cost (Rs) |  |
| CA Lungs | 15.32 | 4.52 | 1.3<br>(USD 0.01) | 15.68 | 4.67 | 0.3<br>(USD 0.00) | 1.6<br>(USD 0.01) |
| COPD | 37.51 | 30.40 | 65<br>(USD 0.46) | 38.39 | 22.37 | 3<br>(USD 0.02) | 68<br>(USD 0.49) |
| CVDs | 35.07 | 36.66 | 78<br>(USD 0.56) | 35.06 | 35.19 | 45<br>(USD 0.32) | 123<br>(USD 0.88) |
| Total Cost | 144<br>(USD 1.03) |  |  | 48<br>(USD 0.34) |  |  | 192<br>(USD 1.37) |

SIR: Smoking Impact Ratio, SAF: Smoking Attributable Fraction

Further analysis showed that majority of burden i.e. Rs. 110 billion (USD 0.78 billion) was related to health care cost (Figure 1).

**Figure 1:**
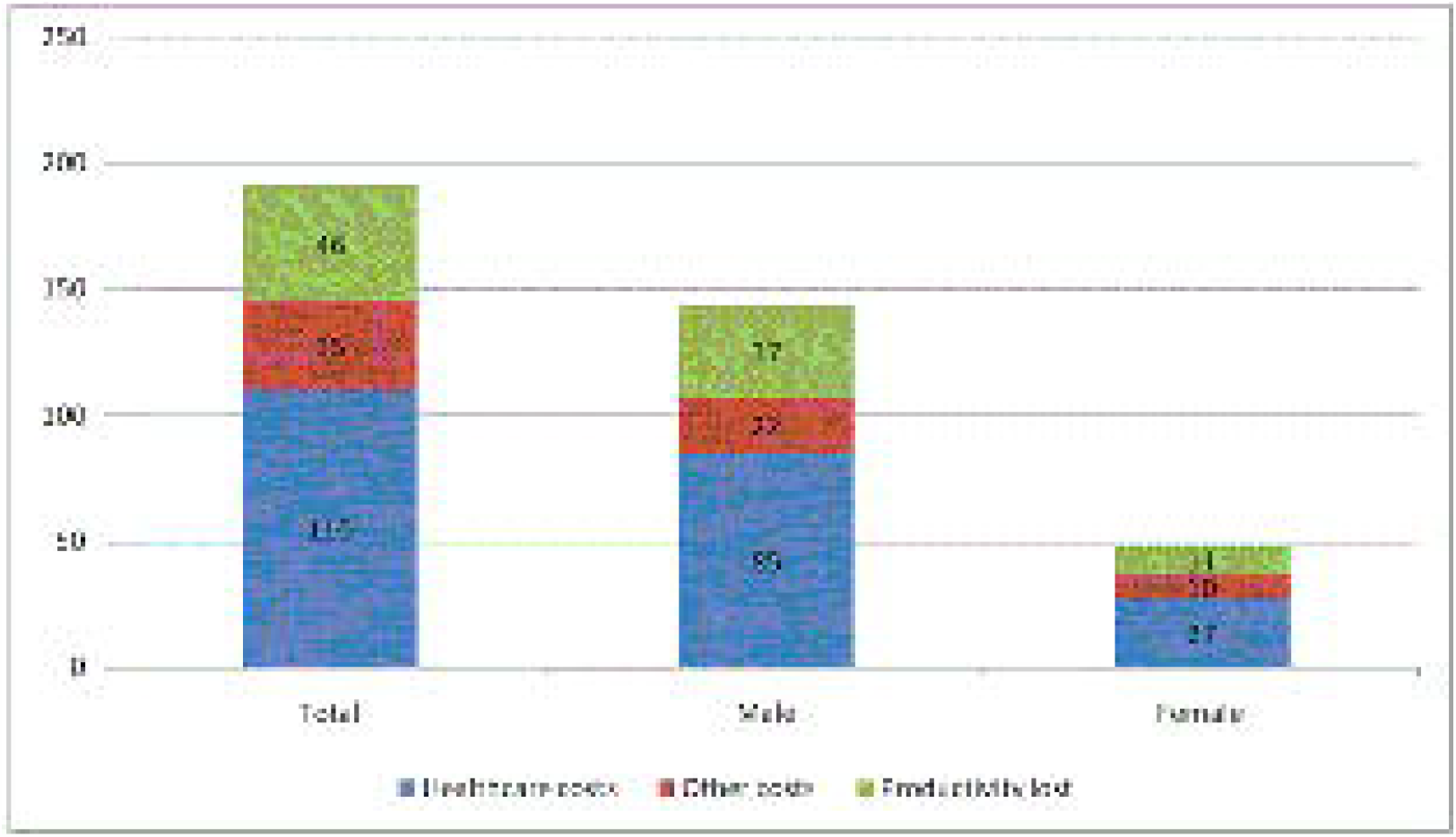
Distribution of total and gender specific smoking attributed economic burden.

## Discussion

Economic burden of tobacco due to its purchases or attributed to illnesses has significant impacts on individuals, families and the country’s economy. Our study revealed that in 2018, tobacco attributed economic burden in Pakistan was Rs.192 billion (USD 1.3 billion) which represented 0.41% of the total GDP (314.58 billion USD) of that year i.e. 2018-19 ^15^.

Overall the economic burden due to tobacco attributed illnesses in Pakistan is almost comparable to the findings from India i.e. $1.7 billion^7^. The GDP lost i.e. 0.41% of Pakistan due to smoking attributed illnesses is higher than reported from Iran (0.26%) ^16^, Korea (0.2%) ^17^ and Sri Lanka (0.15%) ^18^. But it is less than Thailand and Uganda where the smoking attributed burden accounts for 0.7% and 0.5% of the country GDP respectively^19 20^. However, it is almost similar to findings from Taiwan ^21^ where it was 0.4% of the GDP though they included the cost of both smoking and second hand smoke.

The direct cost constituted 60% of the total cost which is contrary to the previous reports in which indirect cost made up the largest portion of the total cost ^22-24^. Similarly, this cost was much higher than reported from Thailand (15%), China (21-25%) and Vietnam (51%) ^19 25 26^. These variations might be due to differences in health care systems. In Pakistan, two-thirds (66%) of the healthcare users access healthcare services from private hospitals and doctors’ fee” constitutes nearly 20% of the out of pocket expenditure ^27 28^. Therefore, the direct cost is high in Pakistan resulting in a huge burden on the patients and families.

For the year 2018-19, the total revenue collected from the tobacco industry in Pakistan was Rs.110 billion (0.74 Billion USD) ^29 30^. It is argued that the tobacco industry contributes to the economy of a country by paying the taxes to the government and creating earnings for tobacco farmers and jobs. However, our findings showed that the total revenue collected from the tobacco industry was 57% of economic burden due to tobacco associated illnesses. Similarly, the total contribution of the tobacco industry to country GDP was equalled to 0.23% while 0.41% of the GDP was lost due to tobacco associated illnesses. Therefore, the contribution of the tobacco industry is marginalized by the fact that the cost bears by the patients and society were much more than the benefits. There is urgent needs to review the cost spend on tobacco associated illnesses and make comparison with the revenue being collected by tobacco industries.

We conclude that tobacco consumption results in a huge economic burden on the patients, families and the country which can be averted by effective implementation of tobacco control policies^31^. One of the best strategies is to increase taxes on tobacco products^32^. It is hoped that the findings of our study will provide sufficient evidence to the policymakers to reassess the current tobacco taxation in Pakistan.

We used robust methods to estimate the economic burden of smoking tobacco on patients and society. However, there are some potential limitations to the study. We have not included the information about smokeless tobacco, second hand smoke and patients suffering from tobacco attributed diseases other than CA lungs, COPD and CVDs. Similarly, other costs like capital invests, salaries etc are not included. Therefore, these findings, if taken alone, might substantially underestimate the total economic burden of tobacco in Pakistan.

## Data Availability

We will share all data once the MS is publihsed

## ACKNOWLEDGEMENTS

We would like to thank all the collaborators for this study. Thanks are due to Mr. Saeed Ahmed Shahid for data entry.

